# Estimating Periodontitis Experience Under Informative Tooth Loss

**DOI:** 10.64898/2026.05.27.26354300

**Authors:** Kym M McCormick, Gina Guzzo, Najith Amarasena

## Abstract

**Background.:** Clinical attachment loss (CAL) is widely used in population oral health research as an indicator of periodontal tissue destruction but can only be measured on teeth present at examination. Retained-dentition summaries may therefore underrepresent cumulative periodontal destruction when tooth loss is partly periodontal in origin.

**Objective.:** To develop and evaluate a framework for estimating periodontitis experience from cross-sectional data by incorporating informative tooth loss under etiological uncertainty.

**Methods.:** Data came from 10,324 dentate adults aged ≥30 years in NHANES 2009–2014 who completed periodontal and glycated hemoglobin (HbA1c) assessments. Expected Periodontitis Experience (EPE) combined observed CAL among retained teeth with the expected periodontal contribution of missing teeth. Published extraction-cause studies informed age-specific weights for missing teeth. EPE was compared with mean CAL and extent CAL ≥4 mm using survey-weighted age patterns and late-life curvature within glycemic groups. HbA1c was used as a theoretically related but distinct external measure.

**Results.:** Periodontal observability declined with age, particularly among adults with diabetes. Conventional observed-site measures showed weaker or declining fitted increases after age 70, whereas EPE retained a more consistently increasing age pattern. Late-life curvature was 0.06, 0.04, and −0.05 across glycemic groups for EPE, compared with −0.47, −0.91, and −2.70 for extent CAL ≥4 mm. EPE also showed the strongest rank correlation with HbA1c (ρ = 0.291 versus 0.238–0.255).

**Conclusions.:** Treating tooth loss as probabilistically informative may provide a more coherent representation of cumulative periodontal destruction in cross-sectional periodontal epidemiology, particularly at older ages.

---

Clinical attachment loss (CAL) is widely used in population oral health research as an indicator of periodontal tissue destruction. CAL estimates the amount of periodontal attachment that has been lost at a measured periodontal site, using the cementoenamel junction as a fixed anatomical reference point ^1^. Because CAL is recorded at individual periodontal sites, it can be summarized in several ways, including as a mean value across sites, as the maximum value observed, or as the percentage of sites exceeding a specified threshold ^2^. These summaries are useful and well established for describing periodontal status among teeth present at examination. In population oral health research, however, CAL-based summaries are also commonly used as person-level indicators of periodontal disease experience in surveillance and epidemiologic analyses ^2,3^. This broader interpretation is more complex because CAL can only be measured where a tooth remains present. When a tooth has already been lost, any attachment loss that occurred on that tooth is no longer observable. Retained-dentition CAL summaries therefore describe periodontal destruction among the teeth available for examination, rather than the full accumulation of periodontal destruction across all teeth that were previously at risk.

This distinction matters because the teeth present at examination may not be a neutral subset of those previously at risk. Tooth retention can partly depend on periodontal history, since teeth with advanced attachment loss have poorer prognoses and are more likely to be extracted or lost ^1,4,5^. Tooth loss therefore changes not only the number of teeth available for examination, but also the type of periodontal history that remains observable. When severely affected teeth are lost preferentially, the sites that would have carried evidence of prior, often severe, destruction are removed from the periodontal record. This creates survivor-conditioned measurement at the tooth level, because periodontal data are available only for teeth that remain present long enough to be measured ^6^. As this process accumulates, CAL summaries may increasingly reflect survivorship of the dentition as well as periodontal severity among retained teeth.

Empirical evidence from NHANES 2009–2014 is consistent with this concern. In prior analyses, CAL summaries showed age-related attenuation, with the strongest attenuation observed for summaries most sensitive to severe attachment loss ^7^. Higher-threshold extent summaries were most affected, whereas broader summaries showed more stable age-related patterns. These patterns raise the possibility that, as periodontal destruction and tooth loss accumulate, conventional retained-dentition summaries may increasingly reflect the periodontal status of teeth that survived to examination rather than cumulative periodontal experience across the dentition previously at risk.

The distinction between retained-dentition periodontal status and cumulative periodontal experience is especially important in studies of ageing and chronic systemic conditions, where cross-sectional periodontal summaries are used to examine associations with health conditions that develop over extended periods ^8–10^. In such studies, the scientific question may concern periodontal destruction accumulated over many years, whereas the summaries available in population datasets are commonly calculated from the retained dentition at a single examination. This creates a mismatch between the intended measurement target and the observed measurement frame, analogous to broader estimand problems in which the empirical quantity available from the data differs from the theoretical target implied by the research question ^11^. The mismatch may be greatest in older adults and in groups with chronic systemic disease, in whom periodontal destruction and tooth loss are more likely to have accumulated. Diabetes provides a particularly relevant setting because of its established associations with both periodontitis and tooth loss ^12,13^. If the most periodontally affected teeth are preferentially lost, attenuation of retained-dentition periodontal summaries may be especially apparent among older adults and adults with diabetes.

This concern is consistent with contemporary periodontal classification, which recognizes that tooth loss attributable to periodontitis contributes to the representation of advanced disease. The 2017 World Workshop classification framework incorporates periodontitis-related tooth loss into staging as an indicator of severity ^14,15^. This reflects the principle that advanced periodontal destruction may not be fully characterized from retained teeth alone. The unresolved problem for population epidemiology is that missing teeth may be observed in cross-sectional survey data, while their reasons for loss are often unavailable. The question is therefore not whether periodontitis-related tooth loss should contribute to measurement, but how missing teeth should be incorporated when their attribution is uncertain.

The present study addresses this problem by estimating Expected Periodontitis Experience (EPE) as a population-level, expectation-based measure of cumulative periodontal destruction under etiological uncertainty. EPE does not attempt to classify individual missing teeth as periodontally lost or reconstruct individual disease histories. Instead, it asks whether missing teeth can contribute probabilistically to an estimate of expected periodontal experience under explicit assumptions about informative tooth loss and uncertain attribution. The construct is therefore best understood as expected cumulative periodontal tissue destruction, not as a measure of current inflammation, lifetime inflammatory activity, systemic-disease risk, or the etiological cause of any individual missing tooth. We examined how EPE behaved relative to conventional CAL summaries as periodontal observability declined with age, including differences in late-life attenuation, and compared their associations with HbA1c as a theoretically related but distinct external measure.

## Method

### Participants

Data were drawn from the 2009–2014 National Health and Nutrition Examination Survey (NHANES), a nationally representative survey of the U.S. population. The primary analytic sample comprised 10,324 dentate adults aged ≥30 years who completed the full-mouth periodontal examination (third molars excluded), demographic interview and laboratory measurement of glycated hemoglobin (HbA1c). Participants had a mean age of 52.02 years (*SD* = 14.25); 5,878 had HbA1c in the normal range, 3,210 in the prediabetes range and 1,236 in the diabetes range according to American Diabetes Association thresholds ^16^. A broader periodontal analytic sample of 10,714 dentate adults with sufficient periodontal examination data, irrespective of HbA1c availability, was used to estimate the age pattern of accumulated missing teeth and the overall age-group descriptive estimates. All analyses were restricted to participants with at least one natural tooth.

This study used publicly available, de-identified NHANES data and was exempt from institutional ethics review. NHANES protocols were approved by the National Centre for Health Statistics Research Ethics Review Board, and all participants provided written informed consent.

### Measures

#### Conventional periodontal summaries

Mean CAL was calculated as the average CAL across all six observed periodontal sites on retained teeth. Extent CAL ≥4 mm was calculated as the percentage of all observed periodontal sites with CAL ≥4 mm. Both summaries were conditioned on retained teeth and observed periodontal sites only.

#### Expected Periodontitis Experience (EPE)

EPE was estimated as a tooth-level measure combining peak observed periodontal destruction among retained teeth with the expected periodontal contribution of missing teeth. For each retained tooth, the maximum CAL across four interproximal sites—mesiofacial, distofacial, mesiolingual and distolingual—was used as the observed value. Mid-facial and mid-lingual sites were excluded so that an isolated non-interproximal value could not determine the tooth-level maximum. The retained-tooth component therefore represents peak interproximal destruction rather than average site-level involvement. EPE was calculated across 28 eligible tooth positions, excluding third molars, placing retained and missing teeth on a common denominator. A schematic of the EPE construction is provided in Supplementary Figure S4.

For missing teeth, an age-specific periodontal-attribution model was estimated from three published extraction-cause studies (Table 1). The fitted incident function, *q*(*a*), represented the probability that an extraction at age was attributed to periodontitis. Because the age at which an already-missing tooth was lost was unknown, was combined with the observed age pattern of accumulated missing teeth in the broader NHANES periodontal sample to derive a cumulative attribution weight for each examination age. Mean missing teeth at age 30 were treated as the initial accumulated stock and weighted by the age-30 incident probability; subsequent annual increments were weighted by the corresponding age-specific probabilities. Full model specification, including the spline form, source weighting, age-band representation, formulae and leave-one-source-out analyses, is provided in Supplementary Appendix A. Selected incident estimates and source-specific fits are reported in Supplementary Table S1 and Supplementary Figure S1. Leave-one-source-out results are reported in Supplementary Table S2 and Supplementary Figure S2, and the estimated age pattern of accumulated missing teeth is shown in Supplementary Figure S3.

**Table 1.** Extraction-attribution schedules.

| Attribution<br>Schedule | Author | Country | Age range<br>(years) | n | Conditional<br>probability |
| --- | --- | --- | --- | --- | --- |
| Schedule A | Chrysanthakopoulos (2011) | Greece | 18–24 | 180 | .022 |
|  |  |  | 25–34 | 446 | .100 |
|  |  |  | 35–44 | 576 | .222 |
|  |  |  | 45–54 | 454 | .370 |
|  |  |  | 55–64 | 529 | .500 |
|  |  |  | 65+ | 277 | .650 |
| Schedule B | Murray et al. (1996) | Canada | 1–12 | 50 | .000 |
|  |  |  | 13–19 | 175 | .000 |
|  |  |  | 20–39 | 535 | .193 |
|  |  |  | 40–59 | 488 | .606 |
|  |  |  | 60+ | 462 | .465 |
| Schedule C | Al-Shammari et al. (2006) | Kuwait | 12–20 | 223 | .009 |
|  |  |  | 21–30 | 506 | .061 |
|  |  |  | 31–40 | 633 | .207 |
|  |  |  | 41–50 | 495 | .451 |
|  |  |  | 51–60 | 592 | .666 |
|  |  |  | 61+ | 334 | .775 |

Each missing tooth position contributed the cumulative attribution weight multiplied by an assigned advanced CAL value. This value was defined as the greater of 10 mm or the participant’s maximum observed tooth-level CAL, calculated using the same four interproximal sites. The assigned value was a calibration parameter rather than an estimate of the true pre-extraction CAL of any missing tooth. Participant-level EPE was calculated as retained-tooth peak burden plus expected missing-tooth burden, divided by 28 eligible tooth positions. Component estimates are reported in Supplementary Table S3, and sensitivity to the 10-mm specification in Supplementary Appendix C. Detailed parameter-sensitivity results are reported in Supplementary Tables S6–S8 and Supplementary Figures S5–S6.

#### HbA1c and Age

HbA1c was analyzed continuously as a theoretically related but distinct external measure and categorized according to American Diabetes Association thresholds for the stratified age-pattern analyses. Age was recorded at examination. Because age is related to periodontal destruction, tooth loss and glycaemia status, age-adjusted models were treated as sensitivity analyses rather than primary models.

### Analytic strategy

Analyses proceeded in three stages. First, we summarized the fitted incident attribution function used to inform EPE. Selected age-specific values of were reported, and leave-one-source-out refits assessed the influence of individual empirical schedules. The incident and cumulative attribution functions used to construct EPE are shown in Figure 1; construction formulae, calibration details and sensitivity analyses are provided in Supplementary Appendix A.

**Figure 1.**
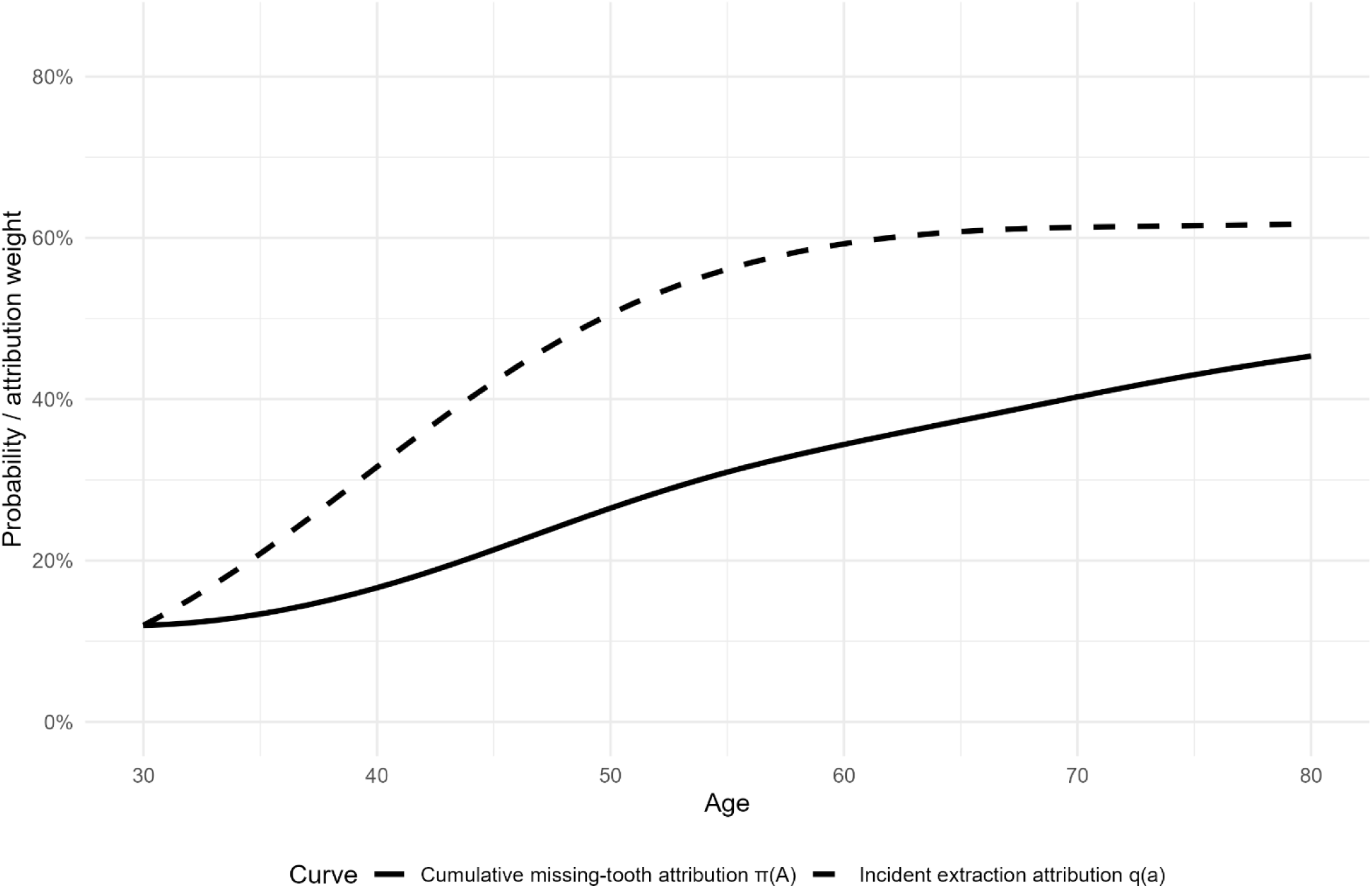
Incident periodontal-attribution function and cumulative missing-tooth attribution weight. The incident attribution function, *q*(*a*) represents the estimated probability that an extraction occurring at age was attributed to periodontitis. The cumulative attribution weight, *π*(*A*) represents the expected periodontal contribution of a tooth already missing at examination age. The cumulative weight combines the incident attribution function with the observed age pattern of accumulated missing teeth and was used to calculate Expected Periodontitis Experience.

Second, we compared EPE with mean CAL, extent CAL ≥4 mm and the percentage of periodontal sites observed as observability declined across age and glycemic categories. Survey-weighted means and 95% confidence intervals were estimated by age group and glycemic category, and survey-weighted quadratic age curves were fitted within glycemic categories. Glycemic categories were used for stratified description rather than prediction. Late-life attenuation was quantified as the fitted change from ages 70–80 years minus that from ages 60– 70 years, with more negative values indicating greater attenuation. Details are provided in Supplementary Appendix B, Section B1.

Third, we compared associations of EPE, mean CAL and extent CAL ≥4 mm with continuous HbA1c, a theoretically related but distinct metabolic measure. Each periodontal measure was standardized using survey-weighted means and standard deviations and analyzed in a separate survey-weighted linear regression model. Coefficients represented the difference in HbA1c percentage points per one-standard-deviation increase in the periodontal measure. Age-adjusted models included linear and quadratic age terms. Design-weighted rank correlations and exploratory joint age-adjusted models were also estimated; details and results are provided in Supplementary Appendix B, Section B2. These analyses provided comparative external evidence and were not intended to evaluate prediction, diagnostic classification or causation.

Confidence intervals for EPE-based estimates reflect NHANES sampling uncertainty conditional on the fitted attribution model and do not incorporate the full uncertainty in the external attribution probabilities. Source dependence was assessed by excluding each extraction-attribution schedule in turn and propagating the refitted function through the cumulative attribution weights. Overall EPE, the expected missing-tooth component, late-life curvature, HbA1c coefficients and rank correlations were then recalculated. Parameter sensitivity was assessed by recalculating EPE using minimum assigned CAL values of 6, 8, 12 and 14 mm, compared with the primary 10-mm specification, and repeating the component, HbA1c, rank-correlation and curvature analyses. These analyses assessed robustness rather than selecting the specification that maximized association with HbA1c. Full methods and results are provided in Supplementary Appendices A and C.

All analyses incorporated the NHANES complex survey design using examination weights, strata and primary sampling units, consistent with NHANES guidance for design-based estimation ^17^. All analyses were conducted in R ^18^ using `survey` for design-based estimation ^19^. Complete analytical code and implementation details are provided in the accompanying repository.

## Results

### Age-related patterns and periodontal observability

Table 2 and Figure 2 summarise age-related patterns in EPE, conventional observed-site periodontal summaries, and periodontal observability. In the broader periodontal analytic sample, the percentage of periodontal sites observed declined markedly with age, from 92.0% among adults aged 30–39 years to 67.9% among those aged 80 years and older. The decline was more pronounced among adults with diabetes, particularly at older ages. Reduced observability was accompanied by attenuation in the age patterning of conventional periodontal summaries. Mean CAL and extent CAL ≥4 mm increased with age, but their fitted age curves showed late-life flattening or downward curvature, most notably among adults with diabetes. Such attenuation is consistent with survivor-conditioned measurement, whereby preferential loss of more severely affected teeth progressively removes evidence of prior destruction from the observable periodontal record. EPE showed less evidence of late-life attenuation than the conventional observed-site summaries. Late-life curvature, defined as the difference between the fitted 10-year changes from ages 70–80 and 60–70 years, was positive among adults with normal glycaemia (0.06 EPE units; 95% CI 0.03, 0.09), close to zero among those with prediabetes (0.04; 95% CI −0.01, 0.08), and modestly negative among those with diabetes (−0.05; 95% CI −0.13, 0.03). By contrast, extent CAL ≥4 mm showed negative curvature in all glycaemic groups, particularly among adults with diabetes, while mean CAL showed its clearest late-life attenuation in the diabetes group. Full estimates are reported in Supplementary Table S4.

**Figure 2.**
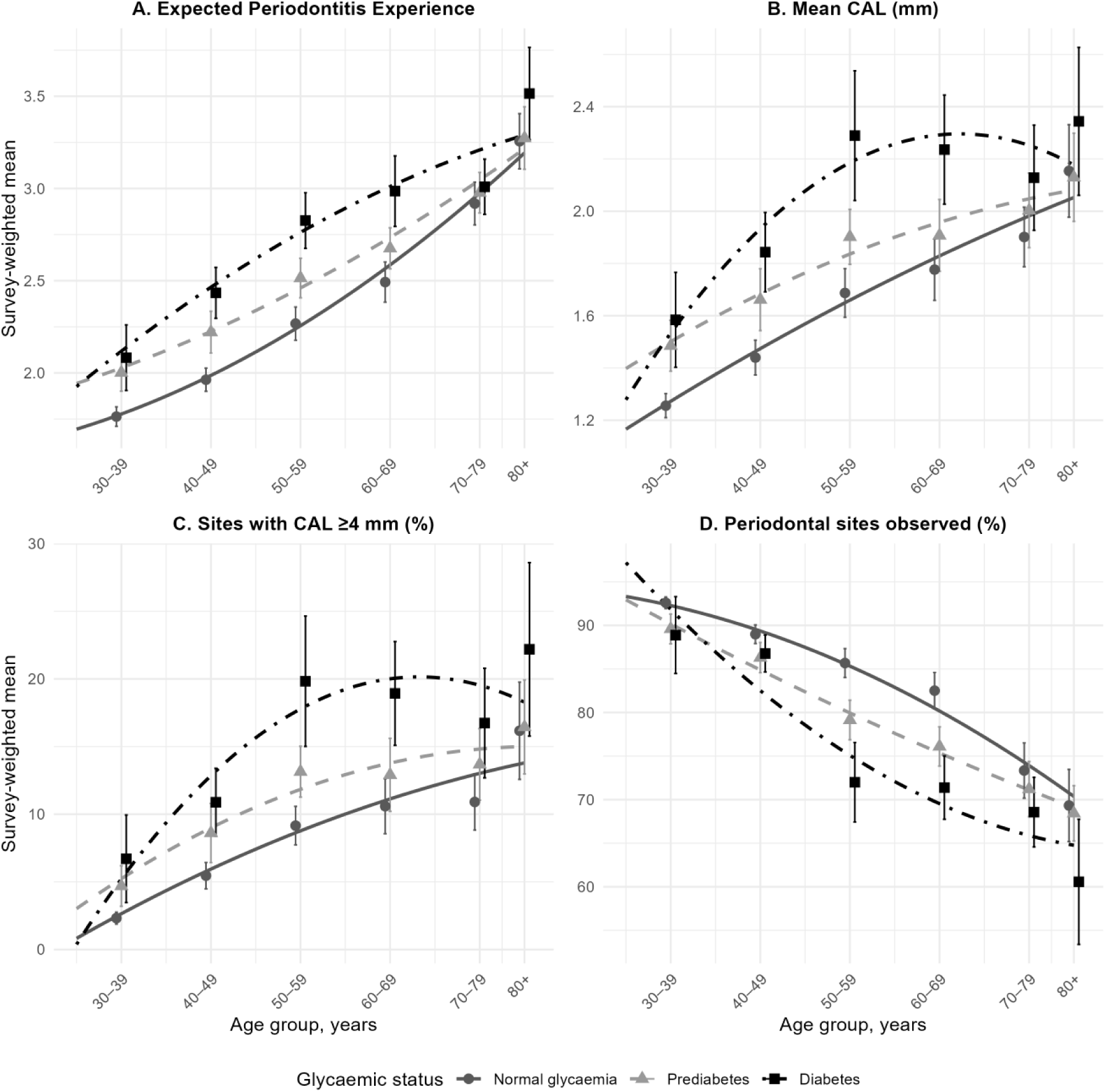
Survey-weighted periodontal measures by age group and glycemic status. Points show survey-weighted age-group means with 95% confidence intervals estimated using the NHANES complex survey design. Points are horizontally offset slightly within age groups to improve visibility. Curves show survey-weighted quadratic fits using continuous age within glycemic-status groups. Panels compare EPE (Panel A) with conventional observed-site measures: mean CAL (Panel B), percentage of observed periodontal sites with CAL ≥ 4 mm (Panel C), and percentage of periodontal sites observed (Panel D). EPE is presented on a mean-tooth scale and incorporates observed attachment-loss burden among retained teeth plus the expected periodontal contribution from missing teeth. CAL = clinical attachment loss; EPE = Expected Periodontitis Experience.

**Table 2.**
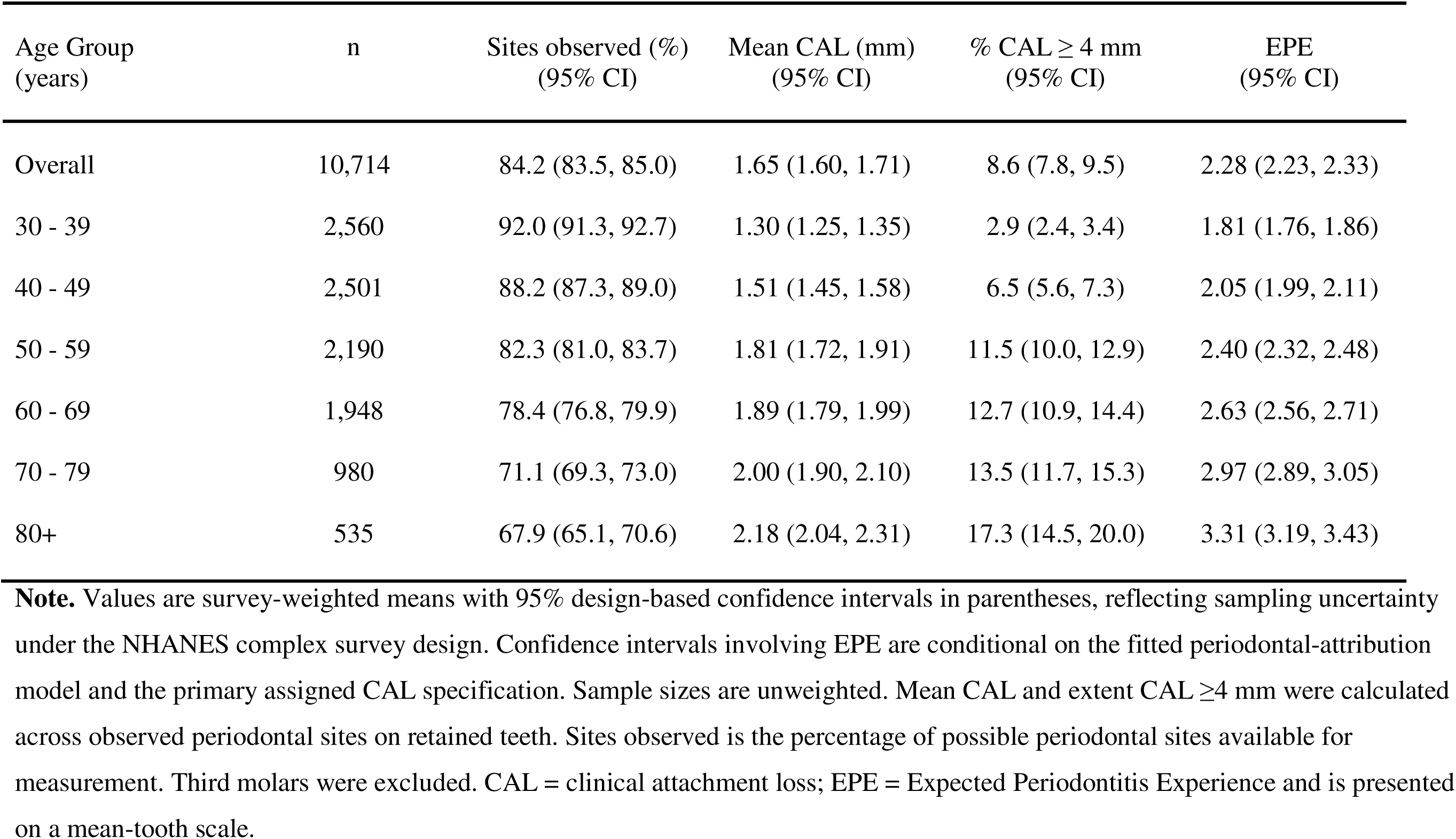
Survey-weighted means and 95% confidence intervals (CIs) for periodontal measures by age group.

### Associations with HbA1c

The association between periodontal measurement and HbA1c was consistently strongest for EPE, followed by mean CAL and extent CAL ≥ 4 mm, although differences between summaries were modest (Table 3). This ordering was unchanged after adjustment for linear and quadratic age. Associations were attenuated by a similar proportion across all summaries. EPE remained positively associated with HbA1c after age adjustment and retained the strongest coefficient among the periodontal summaries.

**Table 3.** Survey-weighted associations between periodontitis measures and HbA1c.

| Periodontal measure | Unadjusted $\beta$ (95% CI) | Age-adjusted $\beta$ (95% CI) | Attenuation after age adjustment |
| --- | --- | --- | --- |
| Expected Periodontitis Experience | 0.201 (0.171, 0.230) | 0.143 (0.112, 0.174) | 28.80% |
| Mean CAL | 0.168 (0.136, 0.201) | 0.122 (0.090, 0.154) | 27.60% |
| Extent CAL $\geq 4$ mm | 0.158 (0.126, 0.189) | 0.114 (0.082, 0.146) | 27.40% |

**Note.** Values are survey-weighted regression coefficients with 95% design-based confidence intervals in parentheses, reflecting sampling uncertainty under the NHANES complex survey design. Confidence intervals involving EPE are conditional on the fitted periodontal-attribution model and the primary assigned CAL specification. Coefficients represent the difference in HbA1c percentage points per one weighted standard-deviation increase in each periodontal measure. Age-adjusted models included linear and quadratic age terms. All associations were statistically significant at $p < .001$ . CAL = clinical attachment loss; EPE = Expected Periodontitis Experience.

Design-weighted rank correlations showed a similar pattern, with EPE again strongest, although the relative order of mean CAL and extent CAL ≥4 mm differed from the regression results. These estimates, along with exploratory joint age-adjusted HbA1c models, are reported in Supplementary Table S5. The joint models were treated as sensitivity analyses because EPE, mean CAL, and extent CAL ≥ 4 mm are correlated and represent overlapping summaries of periodontal burden. The principal age-patterning and HbA1c findings were retained across the source-exclusion and assigned-CAL sensitivity analyses (Supplementary Appendices A and C).

## Discussion

In this study, the consequences of declining periodontal observability differed across the periodontal measures. As fewer sites remained observable with age, mean CAL and extent CAL ≥4 mm showed greater late-life attenuation, particularly among adults with diabetes, whereas EPE retained a more consistently increasing cross-sectional age pattern. EPE also showed modestly stronger associations with HbA1c, and the principal age-patterning and HbA1c findings were retained across alternative attribution sources and assigned CAL values. These results suggest that the way cumulative periodontal experience is operationalized, including whether and how missing tooth positions contribute, can affect both the population pattern produced by a measure and its alignment with an external correlate.

### Measurement target and interpretation

Tooth loss changes not only how much periodontal information remains available, but also which periodontal histories remain represented. A lower or flatter retained-dentition summary may indicate less destruction among the teeth present, but it may also arise because teeth with greater prior destruction have already been lost. Comparisons across age groups or populations therefore depend not only on the CAL values observed, but also on the selection of teeth that remain available for examination ^6,11^.

EPE was developed to address this problem at the population level, not to determine why any individual tooth was lost. By representing all eligible tooth positions under explicit assumptions, it complements summaries restricted to retained teeth. The measures differ in more than their treatment of missing teeth: for retained teeth, EPE uses the maximum CAL across four interproximal sites, whereas mean CAL and extent CAL ≥4 mm summarize all observed sites. Divergence between the measures therefore reflects the combined consequences of these different measurement rules. Even so, their comparison shows whether conclusions about cumulative periodontal experience change when the analytic frame includes all eligible tooth positions rather than only the dentition that survives to examination.

This matters most where tooth loss is common or differs systematically between groups. Populations may vary in access to preventive and restorative care, extraction thresholds and cumulative periodontal destruction, and therefore in how much prior disease remains observable at examination. Restricting measurement to retained teeth may then obscure part of the between-group difference under study. EPE offers a transparent way to retain missing tooth positions in the analytic target while preserving uncertainty about their periodontal attribution.

### External alignment with HbA1c

The HbA1c findings provide limited but supportive external evidence. EPE showed modestly stronger rank and regression associations with HbA1c than the conventional summaries. One interpretation is that incorporating an expected contribution from missing teeth captures variation related to glycemic status that is not fully represented by the conventional retained-site summaries. This pattern is consistent with the intended cumulative measurement target, although it does not establish why EPE showed the stronger association.

The results also do not establish that HbA1c validates EPE. HbA1c reflects glycemic status over the preceding months and is influenced by broader metabolic processes ^20^. Its association with periodontal measures may arise through several causal and non-causal pathways, including shared socioeconomic, behavioral and inflammatory determinants and differential access to preventive and clinical care ^8–10^. The differences between periodontal summaries were also modest and narrowed after age adjustment, indicating that shared age patterning accounted for part of the contrast.

The exploratory joint models require similar caution. EPE, mean CAL and extent CAL ≥4 mm are correlated summaries of overlapping periodontal information. Their joint coefficients are therefore conditional on one another and cannot isolate distinct biological components. The persistence of a positive EPE coefficient is consistent with EPE retaining information associated with HbA1c that is not fully represented by conventional summaries, but it does not establish the source, causal meaning or clinical importance of that information. At most, the HbA1c analyses show that the altered age pattern of EPE was not achieved solely by adding variation unrelated to a theoretically relevant external construct.

### Dependence on modelling assumptions

EPE necessarily depends on modelling assumptions because NHANES does not record when missing teeth were lost or why they were extracted. Its absolute values should therefore be interpreted as model-based expectations rather than recovered periodontal histories. The external extraction-cause studies differed in country, age structure, clinical setting and methods of recording extraction reasons ^21–23^. Leave-one-source-out analyses showed that the incident attribution function was most sensitive to source selection at older ages. This variation was reduced after the incident probabilities were integrated over the observed accumulation of missing teeth, and the principal downstream findings were retained.

Similarly, changing the minimum assigned CAL value altered the level of EPE, as expected, but did not materially change the qualitative age-patterning or HbA1c findings. These sensitivity analyses support the robustness of the finding that EPE showed less late-life flattening than the conventional summaries. They do not establish that any particular EPE value is the true cumulative periodontal burden. Stronger calibration data could change the absolute estimates while leaving the underlying measurement argument intact.

### Scope and limitations

EPE is a population-level expectation, not an individual diagnosis or an attribution of why a particular tooth was lost. It does not estimate lifetime inflammatory exposure, disease incidence, onset or duration, and similar EPE values may arise from different histories of periodontal destruction. Its intended use is in population-level research where cumulative periodontal experience is the target and where substantial tooth loss may have removed evidence that would otherwise contribute to measurement.

The current implementation assigns the same age-specific cumulative attribution weight to every missing eligible tooth within a participant. It therefore does not distinguish tooth-position differences in periodontal vulnerability, restorative history, function or non-periodontal extraction risk. Missing tooth positions are also treated additively, without modelling spatial clustering or within-mouth dependence. Future extensions could incorporate tooth position, anatomical neighborhoods and participant-level risk factors as stronger calibration data become available.

The retained-tooth component also reflects a deliberate measurement choice. Maximum interproximal CAL preserves severe localized destruction that would be diluted by averaging, but it may allow a single site to determine the tooth-level contribution. Alternative tooth-level summaries could combine severity and extent more explicitly. In addition, the present implementation was restricted to dentate adults because a retained-tooth component was required. It therefore does not represent cumulative periodontal experience among edentulous adults, who have experienced the greatest loss of periodontal observability.

Finally, the fitted age curves describe cross-sectional population differences rather than within-person trajectories. Age-related accumulation of missing teeth contributes directly to EPE, and the observed patterns may also reflect cohort effects, historical changes in dental care and differential survival. Longitudinal studies with periodontal measurements before extraction and documented extraction indications are needed to evaluate whether EPE corresponds with subsequently observed periodontal histories. Such data would permit stronger calibration, tooth-specific attribution, examination of within-person change, and eventual extension to edentulous populations.

### Conclusion

The value of EPE lies not in replacing established periodontal indices or claiming recovery of true lifetime disease history. Its contribution is to make the measurement target explicit and to clarify what different measures can, and cannot, represent when tooth loss has reduced periodontal observability. Treating tooth loss as probabilistically informative may provide a more coherent representation of cumulative periodontal destruction in cross-sectional periodontal epidemiology, particularly at older ages.

## Data Availability

All data produced are available online at:

https://wwwn.cdc.gov/nchs/nhanes/search/datapage.aspx?Component=Examination&Cycle=2009-2010

https://wwwn.cdc.gov/nchs/nhanes/search/datapage.aspx?Component=Examination&Cycle=2011-2012

https://wwwn.cdc.gov/nchs/nhanes/search/datapage.aspx?Component=Examination&Cycle=2013-2014

## Acknowledgements

The authors gratefully acknowledge Dr. Sabine Selbach for her thoughtful review of the revised manuscript. Her comments helped improve the manuscript’s clinical precision, readability, and interpretation of periodontal measures.

## Trial registration

Not applicable. This study was a secondary analysis of publicly available, de-identified population survey data and did not involve prospective recruitment, intervention assignment, or clinical trial procedures.

## Data and Code Availability

NHANES data are publicly available at https://www.cdc.gov/nchs/nhanes. Analytical code is publicly available at https://osf.io/x2yma.

## Author Contributions

K.M.M. conceived the study, designed the analytical framework, conducted the data analysis, and drafted the manuscript. Co-authors contributed to conceptual refinement, interpretation of results, and critical revision of the manuscript, and all authors approved the final submitted version.

## Data Availability

The data analysed in this study are publicly available from the National Health and Nutrition Examination Survey (NHANES). Analysis scripts are available at https://osf.io/x2yma.

