## Supplementary Materials for "Estimating Periodontitis Experience Under Informative Tooth Loss"

This supplementary material provides methodological detail and additional analyses that extend the main manuscript without repeating the main Methods section. Appendix A gives the construction and calibration of Expected Periodontitis Experience (EPE), Appendix B provides additional descriptive and external-alignment analyses, and Appendix C reports sensitivity analyses for the assigned CAL value used for missing tooth positions.

### Supplementary Appendix A. Expected Periodontitis Experience

#### A1. Unit of measurement: calculation of EPE at the tooth level

Expected Periodontitis Experience (EPE) was calculated at the level of eligible tooth positions rather than at the level of periodontal sites. This choice was made because EPE is intended to represent cumulative periodontal destruction across the dentition while retaining a fixed denominator. Although clinical attachment loss (CAL) is measured at periodontal sites, site-level reconstruction is not defensible for teeth that are missing at examination. For a missing tooth, the number, location, and severity of previously affected sites are unobserved. Estimating site-specific CAL values for missing teeth would therefore require strong assumptions about the intra-tooth distribution of prior attachment loss and would introduce substantial additional uncertainty.

A tooth-level summary provides a more defensible unit for combining observed retained-tooth burden with the expected periodontal contribution of missing teeth. For each retained natural tooth, the maximum CAL observed across the four interproximal sites—mesiofacial, distofacial, mesiolingual and distolingual—was used as the tooth-level observed periodontal burden. Mid-facial and mid-lingual sites were excluded from this calculation. This differed from the conventional mean CAL, extent CAL ≥4 mm and periodontal-observability summaries, which used all six recorded periodontal sites. The interproximal restriction was used because a maximum is determined by a single site; including mid-facial and mid-lingual sites would therefore allow an isolated non-interproximal value to determine the complete tooth-level contribution. The maximum CAL value was selected because it preserves evidence of advanced localised destruction on retained teeth. Averaging across sites within a tooth would dilute severe localised attachment loss, even though such loss is clinically and conceptually important for representing cumulative periodontal destruction.

Using tooth-level maximum CAL also aligns the retained-tooth component with the missing-tooth component, because both retained teeth and missing tooth positions contribute to the same eligible-tooth scale. The resulting EPE measure is therefore still expressed as an average, but its denominator is not restricted to the teeth that survived to examination. Although not examined in the present analysis, this tooth-level structure also supports future extensions in which cumulative periodontal experience is analyzed by tooth position, intraoral patterning, or network topology.

#### A2. Incident periodontal-attribution function

To represent the expected periodontal contribution of missing teeth, we first estimated an incident periodontal-attribution function, $q\left( a \right)$. This function represents the estimated probability that an extraction occurring at age $a$ was attributed to periodontitis.

The model used age-specific extraction-attribution proportions reported in three published studies. Each observation represented one reported age band within a source schedule and included the proportion of extractions attributed to periodontitis, the reported age-band sample size, and the lower and upper limits of the age band. Closed age bands were represented by their midpoint. For open-ended upper age bands, the upper limit was set to 80 years before calculating the midpoint. This choice aligned the calibration range with the maximum analytically distinguishable age in NHANES, where ages 80 years and older are top-coded as 80.

Let $p_{sj}$ denote the reported periodontal-attribution proportion for age band $j$ in source schedule $s$, $m_{sj}$ its age-band midpoint, and $n_{sj}$ its reported sample size. The incident attribution model was estimated as:

$$logit\{E\left( p_{sj} \right)\}=\beta_{0}+f\left( m_{sj} \right)+\gamma_{s},$$

where $f\left( m_{sj} \right)$ was a natural cubic spline of age with three degrees of freedom and $\gamma_{s}$ represented source-schedule fixed effects. The model was fitted using quasibinomial regression with a logit link. The reported proportions were entered directly as the response, with the corresponding age-band sample sizes used as prior weights. They were not converted into rounded success and failure counts. The quasibinomial specification allowed for dispersion arising from heterogeneity across the empirical source schedules.

Predictions were generated at each integer age from 18 through 80 years for each source schedule. For age $a$, the fitted source-specific prediction was:

$$\hat{q}_{s}\left( a \right)=\mathrm{logit}^{-1}\left[ \hat{\beta}_{0}+\hat{f}\left( a \right)+\hat{\gamma}_{s} \right].$$

The source-adjusted incident attribution function used in the primary analysis was the arithmetic mean of the three source-specific fitted probabilities at each age:

$$\hat{q}\left( a \right)=\frac{1}{S}\sum_{s=1}^{S} \hat{q}_{s}\left( a \right),$$

where $S=3$. Thus, source sample sizes influenced estimation within the quasibinomial model, while the final source-adjusted prediction gave equal weight to each fitted source schedule. The minimum and maximum source-specific fitted probabilities at each age were also retained to describe between-source variation. Selected fitted values are reported in Supplementary Table S1, and the empirical age-band proportions and fitted curves are shown in Supplementary Figure S1.

The incident function describes extractions occurring at a particular age. It was not applied directly to all teeth already missing at a participant’s examination age because the age at which those teeth were lost was unknown. It was instead combined with the observed age pattern of accumulated missing teeth, as described in Section A4.

**Supplementary Table S1.**

*Selected age-specific estimates from the incident periodontal-attribution model*

| Age | Source-adjusted $q\left( a \right)$ | Minimum source-specific $q\left( a \right)$ | Maximum source-specific $q\left( a \right)$ |
| --- | --- | --- | --- |
| 18 | 2.0% | 1.4% | 2.4% |
| 25 | 6.0% | 4.3% | 7.3% |
| 30 | 12.0% | 8.8% | 14.4% |
| 35 | 20.9% | 15.8% | 24.7% |
| 40 | 31.6% | 24.9% | 36.6% |
| 45 | 42.2% | 34.5% | 47.8% |
| 50 | 50.6% | 42.6% | 56.4% |
| 55 | 56.1% | 48.2% | 61.8% |
| 60 | 59.3% | 51.5% | 64.9% |
| 65 | 60.8% | 53.1% | 66.3% |
| 70 | 61.3% | 53.6% | 66.8% |
| 75 | 61.5% | 53.8% | 67.0% |
| 80 | 61.7% | 54.0% | 67.2% |

**Note.** Values are fitted age-specific incident periodontal-attribution probabilities, $q(a)$, representing the estimated probability that an extraction occurring at age $a$ was attributed to periodontitis. The $q$ incident column gives the source-adjusted mean fitted probability across the three empirical attribution schedules. The $q$ min and $q$ max columns show the minimum and maximum fitted probabilities across the source-specific schedules at each age and indicate between-source uncertainty. These incident probabilities were subsequently combined with the observed age-pattern of accumulated missing teeth to derive the cumulative missing-tooth attribution weights used in the EPE calculation; they were not applied directly to all teeth missing at examination. EPE = Expected Periodontitis Experience.

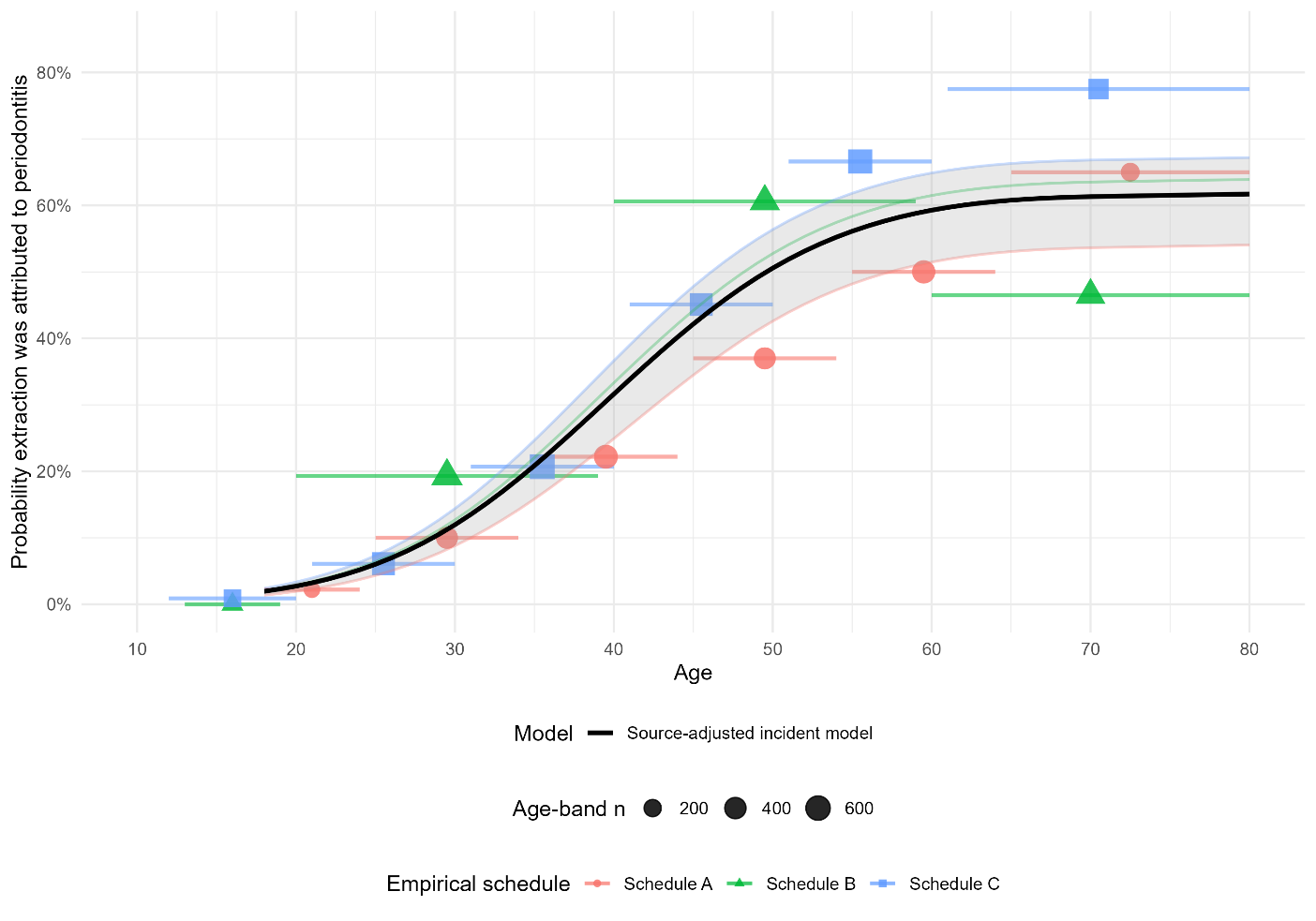

**Supplementary Figure S1. Incident periodontal-attribution model.** Points and horizontal segments show the empirical age-band probabilities that extractions were attributed to periodontitis in the three source schedules presented in Table 1. Point size represents the age-band sample size. The black curve shows the source-adjusted fitted incident attribution function, $q(a)$, estimated using weighted quasibinomial regression with spline terms for age and source schedule. Schedule A = Chrysanthakopoulos (2011); Schedule B = Murray et al. (1996); Schedule C = Al-Shammari et al. (2006).

#### A3. Sensitivity to the empirical extraction-attribution schedules

Because the incident periodontal-attribution function was estimated from three empirical extraction-attribution schedules that differed in country, age-band structure, sample size, and reported attribution probabilities, leave-one-source-out analyses were used to assess dependence on individual sources. Each model was refitted after excluding one schedule, while retaining the same natural-spline, source-effect, weighting, prediction, and aggregation procedures. These analyses served as calibration and robustness checks rather than alternative primary EPE specifications.

The same broad increase in estimated periodontal attribution from age 30 through midlife was retained across all source exclusions (Supplementary Table S2, Panel A; Supplementary Figure S2). At age 30, fitted incident attribution probabilities ranged from 10.0% to 13.8%, and at age 50 they ranged from 46.9% to 57.6%. Greater divergence emerged at older ages. In the full model, the fitted incident attribution probability was 61.3% at age 70 and 61.7% at age 80. When Schedule B was excluded, the corresponding estimates increased to 69.8% and 76.8%; when Schedule C was excluded, they were 54.0% and 53.2%. The older-age portion of the incident attribution function was therefore more sensitive to the available source schedules than the early- and mid-adult portions.

Because incident probabilities were not applied directly to teeth already missing at examination, each leave-one-source-out incident function was subsequently combined with the same observed NHANES age pattern of accumulated missing teeth. The resulting cumulative attribution weights and participant-level EPE values were then recalculated. Variation between source exclusions was attenuated at this downstream stage (Supplementary Table S2, Panel B). At age 80, cumulative missing-tooth attribution weights ranged from 41.9% to 47.7%, compared with 45.3% in the full model. Mean EPE ranged from 2.25 to 2.33 on the mean-tooth scale, and the expected missing-tooth component ranged from 0.43 to 0.50.

The substantive age-patterning and external-alignment results were also retained across source exclusions (Supplementary Table S2, Panel C). Late-life curvature remained positive among adults with normal glycaemia under every specification. Among adults with prediabetes, curvature was positive under every specification, although confidence intervals generally included zero. Among adults with diabetes, curvature remained modestly negative or close to zero, with all confidence intervals including zero. The age-adjusted EPE–HbA1c coefficient ranged from 0.141 to 0.147, and the design-weighted rank correlation with HbA1c ranged from 0.287 to 0.296.

These findings indicate that dependence on individual source schedules was greatest for the older-age portion of the incident attribution function but was substantially reduced after the incident probabilities were integrated over the observed accumulation of missing teeth. The primary model therefore retained the source-adjusted incident attribution function estimated from all three schedules. The principal EPE findings were not materially dependent on any single extraction-attribution schedule. These analyses assess sensitivity to the inclusion of the available empirical schedules but do not represent the full uncertainty in the external calibration evidence.

**Supplementary Table S2**

*Leave-one-source-out sensitivity of the incident periodontal-attribution function and downstream EPE results*

**Panel A. Incident periodontal-attribution probabilities**

| **Model** | **Age 30 years** | **Age 40 years** | **Age 50 years** | **Age 60 years** | **Age 70 years** | **Age 80 years** |
| --- | --- | --- | --- | --- | --- | --- |
| All schedules | 12.0% | 31.6% | 50.6% | 59.3% | 61.3% | 61.7% |
| Without Schedule A | 13.1% | 36.9% | 57.6% | 63.3% | 60.9% | 57.0% |
| Without Schedule B | 10.0% | 27.5% | 46.9% | 60.8% | 69.8% | 76.8% |
| Without Schedule C | 13.8% | 33.0% | 47.6% | 53.4% | 54.0% | 53.2% |

**Panel B. Cumulative attribution weights and EPE components**

| **Model** | **Cumulative attribution weight, age 60** | **Cumulative attribution weight, age 70** | **Cumulative attribution weight, age 80** | **Mean EPE (95% CI)** | **Expected missing-tooth component (95% CI)** |
| --- | --- | --- | --- | --- | --- |
| All schedules | 34.4% | 40.3% | 45.3% | 2.28 (2.23, 2.33) | 0.45 (0.43, 0.48) |
| Without Schedule A | 38.7% | 44.0% | 47.5% | 2.33 (2.28, 2.38) | 0.50 (0.47, 0.53) |
| Without Schedule B | 31.8% | 39.5% | 47.7% | 2.25 (2.21, 2.30) | 0.43 (0.40, 0.45) |
| Without Schedule C | 33.7% | 38.3% | 41.9% | 2.27 (2.23, 2.32) | 0.45 (0.42, 0.47) |

**Panel C. Late-life curvature and HbA1c alignment**

| **Model** | **Normal glycaemia curvature (95% CI)** | **Prediabetes curvature (95% CI)** | **Diabetes curvature (95% CI)** | **Age-adjusted HbA1c β (95% CI)** | **Rank correlation with HbA1c** |
| --- | --- | --- | --- | --- | --- |
| All schedules | 0.060 (0.034, 0.086) | 0.038 (-0.008, 0.084) | -0.049 (-0.126, 0.028) | 0.143 (0.112, 0.174) | 0.291 |
| Without Schedule A | 0.055 (0.028, 0.082) | 0.021 (-0.027, 0.069) | -0.077 (-0.157, 0.004) | 0.147 (0.115, 0.179) | 0.296 |
| Without Schedule B | 0.080 (0.054, 0.105) | 0.069 (0.024, 0.115) | -0.007 (-0.083, 0.069) | 0.141 (0.109, 0.172) | 0.290 |
| Without Schedule C | 0.045 (0.019, 0.071) | 0.022 (-0.023, 0.067) | -0.067 (-0.143, 0.008) | 0.142 (0.111, 0.173) | 0.287 |

**Note.** **Panel A** reports fitted age-specific incident periodontal-attribution probabilities, ($q(a)$), representing the estimated probability that an extraction occurring at a given age was attributed to periodontitis. **Panel B** reports the cumulative missing-tooth attribution weights obtained after combining each incident function with the observed age pattern of accumulated missing teeth in NHANES. Mean EPE and the expected missing-tooth component are presented on the mean-tooth scale. **Panel C** reports late-life curvature calculated as the fitted change from ages 70–80 years minus the fitted change from ages 60–70 years. Positive values indicate a larger fitted increase during ages 70–80 years; negative values indicate attenuation, flattening, or downward curvature. HbA1c coefficients represent the difference in HbA1c percentage points associated with a one survey-weighted standard-deviation increase in EPE, adjusted for linear and quadratic age. Rank correlations are design-weighted correlations between ranked EPE and ranked HbA1c. Confidence intervals are design-based and conditional on the refitted attribution model and the primary 10-mm assigned CAL specification; they do not incorporate the full uncertainty in the external calibration evidence. The “All schedules” model includes all three empirical extraction-attribution schedules. EPE = Expected Periodontitis Experience; CAL = clinical attachment loss; Schedule A = Chrysanthakopoulos (2011); Schedule B = Murray et al. (1996); Schedule C = Al-Shammari et al. (2006).

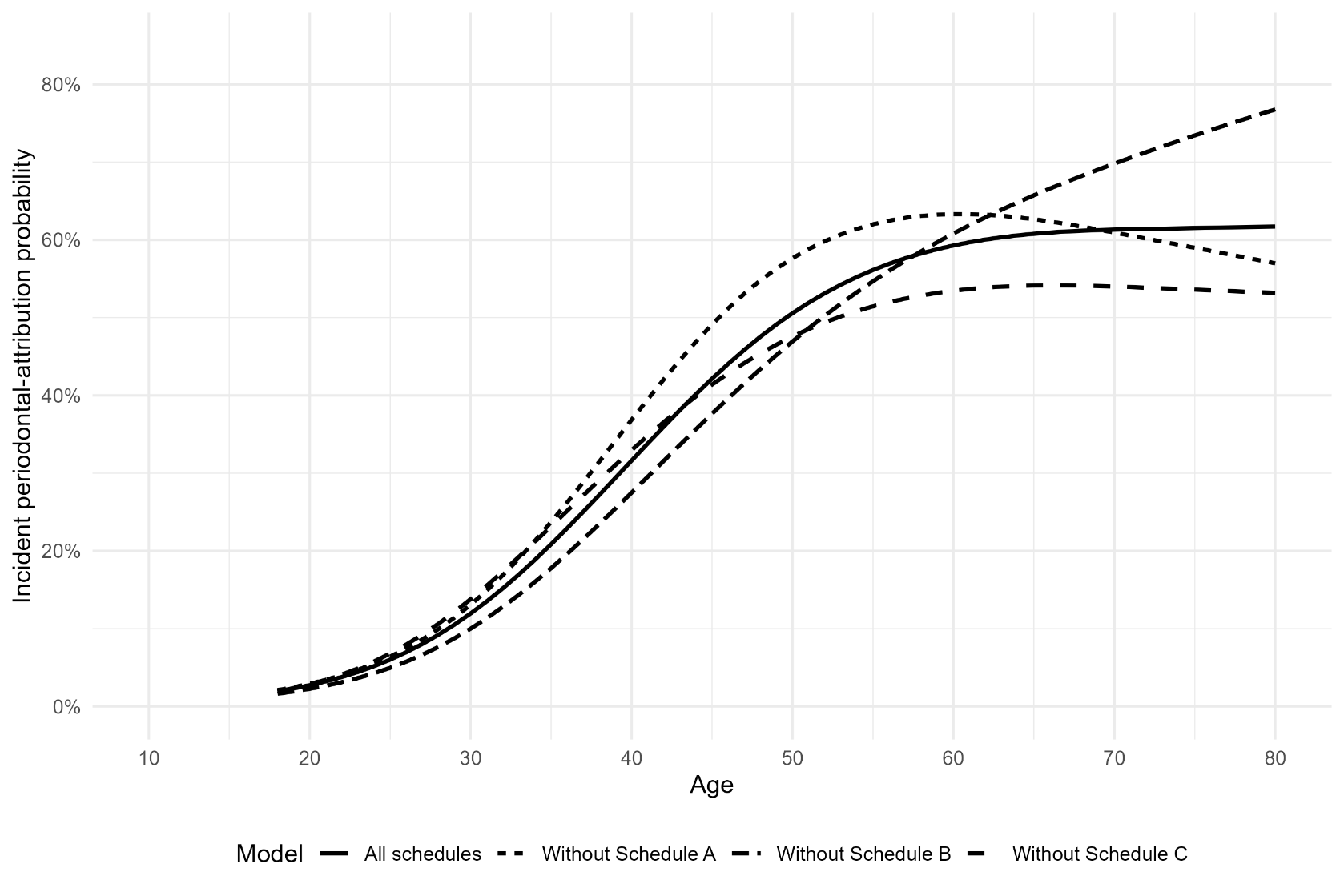

**Supplementary Figure S2.** **Leave-one-source-out sensitivity of the incident periodontal-attribution model.** Curves show fitted incident attribution functions estimated using all three empirical source schedules and after excluding one schedule at a time. The analysis assesses the influence of individual source studies on the fitted incident attribution function, particularly at older ages where the schedules diverged most. Schedule A = Chrysanthakopoulos (2011); Schedule B = Murray et al. (1996); Schedule C = Al-Shammari et al. (2006). Full source schedules are presented in Table 1.

#### A4. Cumulative missing-tooth attribution weight

The incident attribution function, $q\left( a \right)$, is appropriate for an extraction occurring at a specified age. It cannot be applied directly to a tooth already missing at examination because the age at extraction is not observed. A tooth missing at age 70, for example, may have been lost at age 35, 50, 65, or another earlier age. We therefore combined the incident attribution function with the observed age pattern of accumulated missing teeth in NHANES to derive a cumulative missing-tooth attribution weight, $\pi\left( A \right)$, for examination age $A$.

##### A4.1. Estimation of the missing-tooth accumulation curve

The broader periodontal analytic sample of 10,714 dentate adults was used to estimate the number of eligible tooth positions already missing at each age (Supplementary Figure S3). Third molars were excluded, leaving 28 eligible tooth positions. For each integer age from 30 through 80 years, the survey-weighted mean number of missing tooth positions was estimated using the NHANES examination weights, strata, and primary sampling units.

The survey-weighted age-specific means were then smoothed using an ordinary linear regression in which mean missing teeth was modelled as a natural cubic spline of age with four degrees of freedom:

$$E\{M\left( a \right)\}=\alpha_{0}+g\left( a \right),$$

where $M\left( a \right)$ denotes the mean number of missing eligible tooth positions at age $a$, and $g\left( a \right)$ is a natural cubic spline with four degrees of freedom. The regression was weighted by the unweighted number of participants contributing to each single-year age estimate. Predictions were generated at each integer age from 30 through 80 years and constrained to the possible range of zero to 28 missing tooth positions.

Because the intended quantity was accumulated tooth loss, the smoothed sequence was required to be non-decreasing with age. A weighted pooled-adjacent-violators algorithm was applied to the smoothed predictions, using the number of participants at each age as the weights. This produced the ordered accumulation curve, $\hat{M}^{*}\left( a \right)$.

NHANES records participants aged 80 years or older as age 80. The accumulation curve and attribution functions were therefore estimated only through age 80. Participants in the 80-years-and-older group received the cumulative attribution weight evaluated at age 80; no extrapolation beyond the top-coded age was undertaken.

##### A4.2. Annual increments in accumulated missing teeth

The estimated increment in accumulated missing teeth at age $a$ was calculated as the first difference of the ordered accumulation curve:

$$\Delta M\left( a \right)=\hat{M}^{*}\left( a \right)-\hat{M}^{*}\left( a-1 \right).$$

Any negative numerical differences were set to zero, consistent with the non-decreasing accumulation constraint. Because the NHANES periodontal analytic sample began at age 30, the mean number of missing tooth positions observed at age 30 was treated as the initial accumulated stock:

$$\Delta M\left( 30 \right)=\hat{M}^{*}\left( 30 \right).$$

This initial stock was weighted using the age-30 incident periodontal-attribution probability. For ages 31 through 80, each annual increment was weighted by the corresponding fitted incident probability, $\hat{q}\left( a \right)$.

##### A4.3. Calculation of the cumulative attribution weight

For examination age $A$, the cumulative missing-tooth attribution weight was calculated as:

$$\pi\left( A \right)=\frac{\sum_{a=30}^{A} \hat{q}\left( a \right)\Delta M\left( a \right)}{\sum_{a=30}^{A} \Delta M\left( a \right)}.$$

The numerator represents the expected periodontal component of missing-tooth accumulation through age $A$, while the denominator represents total accumulated missing tooth positions through that age. Thus, $\pi\left( A \right)$ is the weighted average of the incident periodontal-attribution probabilities over the ages at which missing teeth accumulated in the population.

The raw cumulative attribution sequence was constrained to be non-decreasing by applying the cumulative maximum:

$$\hat{\pi}^{*}\left( A \right)=\max_{30\leq a\leq A}\hat{\pi}\left( a \right).$$

This restriction was included to preserve the intended interpretation of $\pi\left( A \right)$ as an accumulated attribution weight for teeth already missing by examination age. The unconstrained cumulative estimates were already non-decreasing, so this step did not alter the primary fitted values.

For participant-level EPE calculations, cumulative attribution weights were obtained by linear interpolation over the integer-age lookup table. Ages below or above the fitted range were bounded at the nearest available value. In practice, the analytic sample was restricted to ages 30 through 80 because of the NHANES eligibility range and age top-coding.

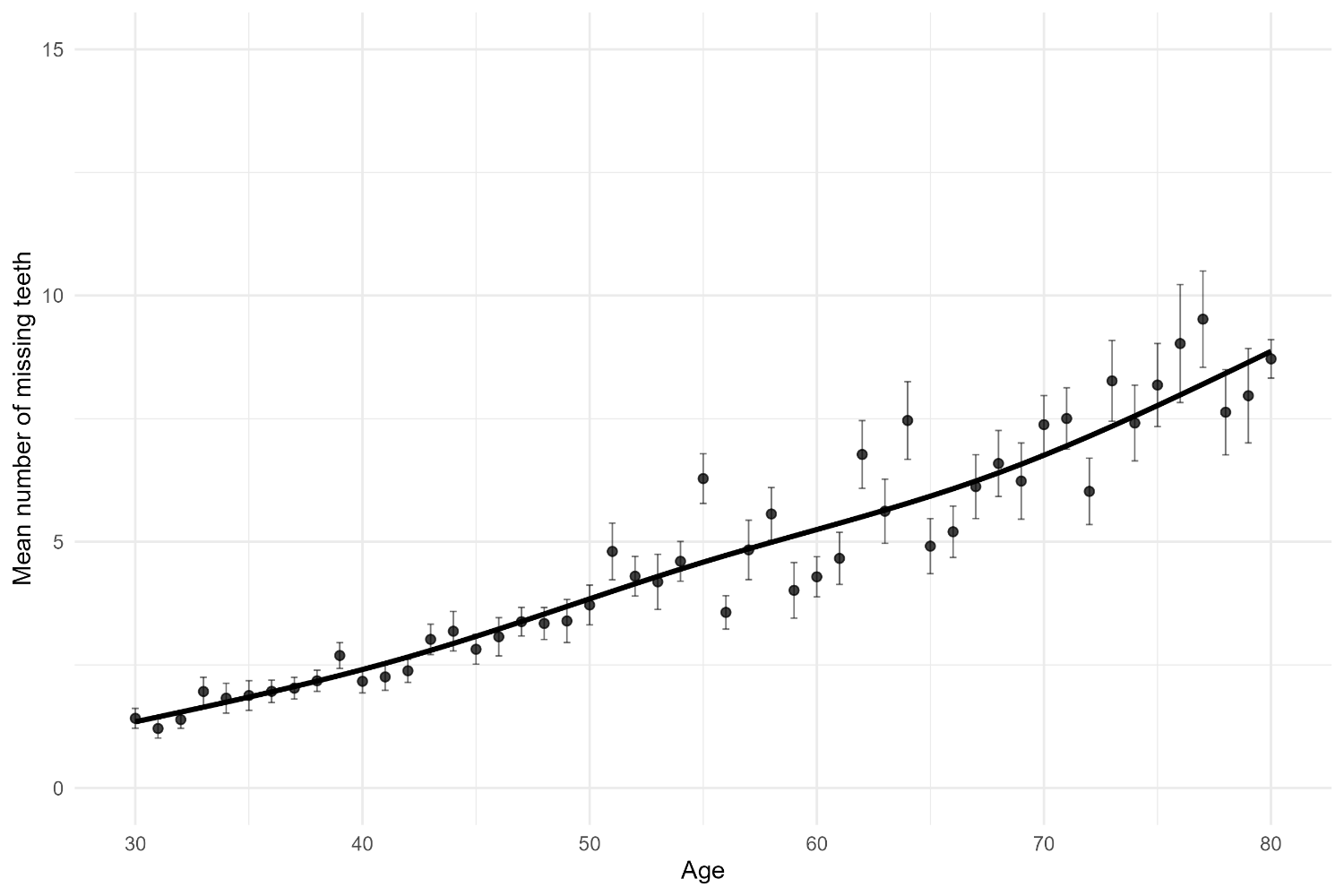

**Supplementary Figure S3. Estimated accumulation of missing teeth by age**. Points show survey-weighted age-specific mean numbers of missing eligible tooth positions, with standard errors, among 10,714 dentate adults aged ≥30 years in the broader NHANES 2009–2014 periodontal analytic sample, irrespective of HbA1c availability. Third molars were excluded. The fitted line shows the smoothed non-decreasing accumulation curve used to estimate the age-specific increments in missing teeth. These increments were used to weight the incident periodontal-attribution probabilities when deriving the cumulative missing-tooth attribution weights used in EPE.

#### A5. Expected contribution from missing teeth

Missing tooth positions were not classified as periodontally lost or not periodontally lost. Instead, each missing tooth position contributed an expected periodontal-experience value. This expected value combined two components: the cumulative missing-tooth attribution weight and an assigned advanced CAL value. The cumulative missing-tooth attribution weight, $\pi(Aᵢ)$, represented the expected probability that a tooth already missing at participant i’s examination age reflected prior periodontal loss. The assigned advanced CAL value, $Bᵢ$, represented the underlying level of severe periodontal destruction assigned to missing tooth positions contributing to EPE.

For participant *i*, the expected contribution for each missing tooth position was calculated as:

$$\text{Expected missing-tooth contribution}ᵢ = \pi(Aᵢ) \times Bᵢ$$

where $\pi(Aᵢ)$ is the cumulative missing-tooth attribution weight for participant i at examination age $Aᵢ$, and $Bᵢ$ is the assigned advanced CAL value for missing tooth positions in that participant.

The assigned advanced CAL value was defined as the greater of 10 mm or the participant’s maximum observed tooth-level CAL:

$$Bᵢ = max(10, \text{maxCAL}ᵢ)$$

where maxCALᵢ is the largest tooth-level maximum CAL observed among retained teeth for participant *i*. This rule was not intended to estimate the true pre-extraction CAL of any individual missing tooth. Rather, it provided a conservative advanced-destruction value for the expected missing-tooth component. The participant-specific maximum rule ensured internal consistency, so that a missing tooth position contributing to EPE was not assigned a lower underlying burden than the most severely affected tooth still observed in the same participant.

Note that the participant maximum used in this rule was the maximum of the retained-tooth values calculated across the four interproximal sites. The same interproximal site definition therefore informed both the observed retained-tooth component and the participant-specific advanced CAL assignment used for missing tooth positions.

Sensitivity to the 10-mm primary specification is reported in Supplementary Appendix C.

#### A6. Participant-level EPE

Participant-level EPE was calculated across all eligible tooth positions, excluding third molars. Retained tooth positions contributed their observed tooth-level maximum CAL values, calculated across the four interproximal sites. Missing tooth positions contributed their expected missing-tooth values, calculated as the cumulative missing-tooth attribution weight multiplied by the assigned advanced CAL value.

EPE was expressed as the mean of retained-tooth and expected missing-tooth contributions across the eligible dentition:

$$EPEᵢ = \frac{\left[ \Sigmaⱼ\in Rᵢ \text{MaxCAL}ᵢⱼ + \Sigmaⱼ\in Mᵢ \pi(Aᵢ)Bᵢ \right]}{Nᵢ}$$

where $Rᵢ$ denotes retained eligible tooth positions for participant i, Mᵢ denotes missing eligible tooth positions, MaxCALᵢⱼ is the maximum observed CAL for retained tooth j, $\pi(Aᵢ)$ is the cumulative missing-tooth attribution weight at participant age Aᵢ, Bᵢ is the assigned advanced CAL value, and $Nᵢ$ is the total number of eligible tooth positions.

Because $\pi(Aᵢ)$ and $Bᵢ$are participant-level quantities, the missing-tooth component can equivalently be written as:

$$\sum_{j\in M_{i}} \pi\left( A_{i} \right)B_{i}=\mid M_{i}\mid\pi\left( A_{i} \right)B_{i}$$

where $\mid M_{i}\mid$ is the number of missing eligible tooth positions for participant $i$.

Thus, EPE is a mean-tooth measure. Its key distinction from conventional retained-dentition summaries is that the denominator is the eligible dentition rather than only the teeth present at examination. Retained teeth contribute observed periodontal burden, while missing tooth positions contribute expected periodontal burden under uncertainty. EPE therefore remains an average, but the average is taken over the dentition at risk rather than only over the surviving dentition.

To make the contribution of each component transparent, Supplementary Table S3 summarises the retained-tooth and missing-tooth components of EPE by age group. The table reports the mean number of retained and missing eligible tooth positions, the cumulative missing-tooth attribution weight, the assigned advanced CAL value for missing teeth, the expected contribution per missing tooth position, and the observed and expected components after scaling by the eligible dentition denominator. This table is descriptive and was used to show how the EPE score is assembled, rather than to test a separate hypothesis.

A schematic illustrating the construction of EPE is given in Supplementary Figure S4.

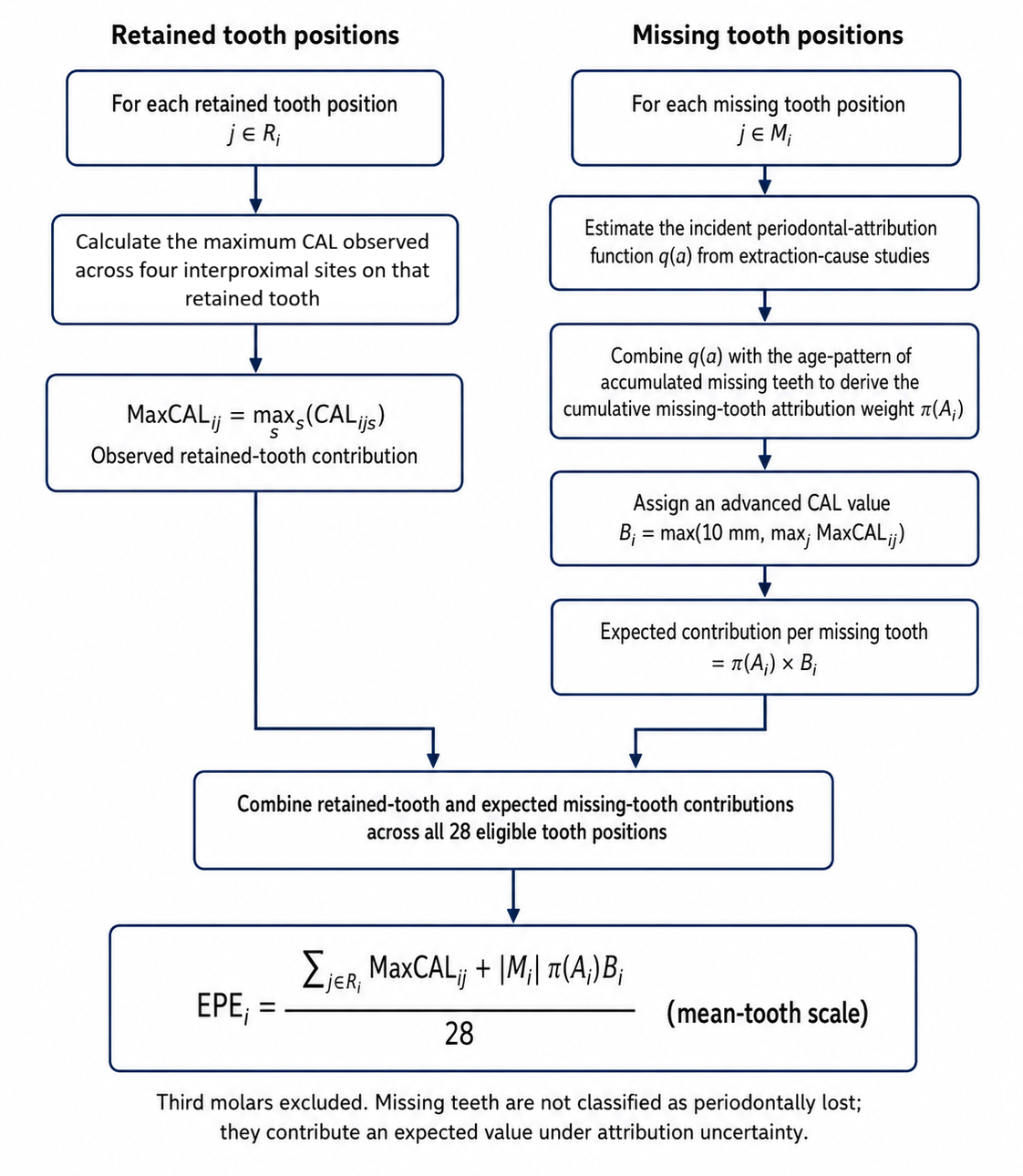

**Supplementary Figure S4. Schematic construction of Expected Periodontitis Experience (EPE).** For retained tooth positions, the observed contribution was defined as the maximum clinical attachment loss (CAL) recorded across the four interproximal sites on that tooth; mid-facial and mid-lingual sites were excluded. For missing tooth positions, the expected contribution was calculated as the cumulative missing-tooth attribution weight multiplied by the assigned advanced CAL value. Retained-tooth and expected missing-tooth contributions were summed across 28 eligible tooth positions, excluding third molars, and divided by 28 to express EPE on a mean-tooth scale. Missing teeth were not classified as periodontally lost; instead, they contributed expected values under periodontal-attribution uncertainty.

**Supplementary Table S3**

*Components contributing to EPE by age group*

| Age Group | n | Retained teeth | Missing tooth positions | Cumulative missing-tooth attribution weight (%) | Assigned advanced CAL for missing teeth (mm) | Expected contribution per missing tooth position (mm) | Observed retained-tooth component | Expected missing-tooth component |
| --- | --- | --- | --- | --- | --- | --- | --- | --- |
| Overall | 10714 | 24.0 (23.8, 24.2) | 4.0 (3.8, 4.2) | 26.2 (25.8, 26.6) | 10.02 (10.02, 10.03) | 2.63 (2.59, 2.67) | 1.83 (1.79, 1.86) | 0.45 (0.43, 0.48) |
| 30-39 | 2560 | 26.1 (25.9, 26.3) | 1.9 (1.7, 2.1) | 13.5 (13.4, 13.5) | 10.00 (10.00, 10.00) | 1.35 (1.34, 1.35) | 1.72 (1.66, 1.77) | 0.09 (0.08, 0.10) |
| 40-49 | 2501 | 25.1 (24.9, 25.3) | 2.9 (2.7, 3.1) | 20.9 (20.7, 21.0) | 10.01 (10.00, 10.01) | 2.09 (2.07, 2.11) | 1.83 (1.78, 1.88) | 0.22 (0.20, 0.24) |
| 50-59 | 2190 | 23.5 (23.1, 23.8) | 4.5 (4.2, 4.9) | 30.3 (30.1, 30.4) | 10.04 (10.03, 10.06) | 3.04 (3.02, 3.06) | 1.90 (1.84, 1.97) | 0.50 (0.45, 0.54) |
| 60-69 | 1948 | 22.3 (21.9, 22.7) | 5.7 (5.3, 6.1) | 36.7 (36.6, 36.9) | 10.04 (10.03, 10.06) | 3.69 (3.68, 3.70) | 1.88 (1.82, 1.93) | 0.76 (0.70, 0.81) |
| 70-79 | 980 | 20.3 (19.7, 20.8) | 7.7 (7.2, 8.3) | 42.3 (42.2, 42.5) | 10.02 (10.01, 10.04) | 4.24 (4.23, 4.26) | 1.79 (1.73, 1.85) | 1.18 (1.10, 1.26) |
| 80+ | 535 | 19.3 (18.5, 20.0) | 8.7 (8.0, 9.5) | 45.3 (45.3, 45.3) | 10.04 (10.01, 10.07) | 4.55 (4.54, 4.57) | 1.89 (1.79, 1.98) | 1.42 (1.29, 1.55) |

**Note.** Values are survey-weighted means with 95% design-based confidence intervals in parentheses. Confidence intervals for the expected missing-tooth component are conditional on the fitted periodontal-attribution model and the primary assigned CAL specification. Component contributions are expressed on the mean-tooth scale used for EPE. The observed retained-tooth component equals the sum of retained-tooth maximum CAL values divided by the number of eligible tooth positions. The expected missing-tooth component equals the expected missing-tooth burden divided by the number of eligible tooth positions. EPE is obtained by summing these two components; overall EPE estimates are reported in Table 2. Sample sizes are unweighted. This table uses the broader periodontal analytic sample ($n=10,714$), rather than the HbA1c subsample. Third molars were excluded. CAL = clinical attachment loss.

### Supplementary Appendix B. Additional descriptive and HbA1c association analyses

This appendix provides additional estimates supporting the descriptive age-patterning, late-life curvature, and HbA1c construct-alignment. These analyses were intended to clarify how Expected Periodontitis Experience (EPE) behaved relative to conventional observed-site periodontal summaries, particularly in older adults and in groups where periodontal observability was reduced. Note that reported confidence intervals are design-based and reflect sampling uncertainty under the NHANES complex survey design. For EPE estimates, they are conditional on the fitted periodontal-attribution model and the specified assigned CAL value; uncertainty in these calibration components was examined separately through leave-one-source-out and parameter sensitivity analyses.

#### B1. Late-life curvature

Supplementary Table S4 reports fitted changes in periodontal summaries from ages 60–70 years and 70–80 years, estimated from survey-weighted quadratic regression models within glycemic-status groups. The late-life curvature statistic was calculated as the fitted change from 70–80 years minus the fitted change from 60–70 years. Negative values therefore indicate attenuation, flattening, or downward curvature in later life.

Across glycemic-status groups, conventional observed-site summaries showed more evidence of late-life attenuation than EPE. Among adults with normal glycaemia, EPE showed slightly greater fitted increase from 70–80 years than from 60–70 years, whereas extent CAL ≥4 mm showed negative late-life curvature. Among adults with prediabetes, EPE again showed little evidence of attenuation, while extent CAL ≥4 mm showed a larger reduction in fitted increase from 70–80 years. The strongest contrast was observed among adults with diabetes. In this group, EPE showed modest attenuation, whereas extent CAL ≥4 mm declined from 70–80 years and mean CAL also showed negative late-life curvature. These results support the interpretation that conventional observed-site summaries are more vulnerable to late-life flattening when periodontal observability is reduced.

.

**Supplementary Table S4.**

*Fitted late-life change and curvature in periodontal summaries from ages 60–80 years, by glycaemia status.*

| Glycemic status | Measure | $\triangle60-70$ years | $\triangle70-80$ years | Late-life Curvature (95% CI) |
| --- | --- | --- | --- | --- |
| Normal | EPE | 0.36 | 0.42 | 0.06 (0.03, 0.09) |
|  | % CAL $\geq$4 mm | 2.13 | 1.66 | -0.47 (-0.92, -0.02) |
|  | Mean CAL | 0.16 | 0.15 | -0.02 (-0.05, 0.01) |
| Prediabetes | EPE | 0.29 | 0.33 | 0.04 (-0.01, 0.08) |
|  | % CAL $\geq$4 mm | 1.49 | 0.58 | -0.91 (-1.73, -0.08) |
|  | Mean CAL | 0.11 | 0.07 | -0.03 (-0.07, 0.01) |
| Diabetes | EPE | 0.22 | 0.17 | -0.05 (-0.13, 0.03) |
|  | % CAL $\geq$4 mm | 0.88 | -1.83 | -2.70 (-4.55, -0.86) |
|  | Mean CAL | 0.03 | -0.11 | -0.15 (-0.24, -0.05) |

**Note.** Values are model-based fitted changes from survey-weighted quadratic regression models estimated separately within glycaemia-status groups. The 60–70 and 70–80 columns represent the fitted change in each periodontal measure over those age intervals. Late-life curvature was calculated as the fitted change from ages 70–80 years minus the fitted change from ages 60–70 years. Positive values indicate a larger fitted increase from 70–80 than from 60–70 years; negative values indicate attenuation, flattening, or downward curvature in later life. Confidence intervals are design-based and conditional on the fitted periodontal-attribution model and the assigned CAL specification shown in each row. They do not incorporate uncertainty in the external attribution calibration. EPE is presented on a mean-tooth scale. Mean CAL is expressed in millimeters, % CAL ≥4 mm is expressed in percentage points.

CAL = clinical attachment loss; EPE = Expected Periodontitis Experience.

#### B2. Rank correlations and exploratory joint HbA1c models

Supplementary Table S5 reports design-weighted rank correlations between each periodontal measure and HbA1c, along with exploratory joint age-adjusted HbA1c models. The rank correlations describe monotonic associations between periodontal summaries and HbA1c. EPE showed the strongest rank correlation with HbA1c, followed by extent CAL ≥4 mm and mean CAL. Periodontal sites observed was negatively correlated with HbA1c, indicating that lower periodontal observability was associated with higher HbA1c.

The exploratory joint age-adjusted model included EPE, mean CAL, extent CAL ≥4 mm, linear age, and quadratic age simultaneously. In this model, EPE retained a positive association with HbA1c, whereas mean CAL and extent CAL ≥4 mm showed attenuated and imprecise conditional associations. These coefficients should be interpreted cautiously because the periodontal summaries are correlated and represent overlapping summaries of periodontal burden. The joint model was therefore treated as a sensitivity analysis rather than primary evidence. Its purpose was to assess whether EPE retained information not fully represented by conventional retained-dentition summaries when the summaries were considered together.

**Supplementary Table S5.**

*Rank correlations with HbA1c and exploratory joint age-adjusted HbA1c models.*

| Measure | $\rho$ with HbA1c | Joint age-adjusted  $\beta$ (95% CI) |
| --- | --- | --- |
| Expected Periodontitis Experience | 0.291 | 0.138 (0.086, 0.191) |
| Mean CAL | 0.238 | -0.037 (-0.123, 0.049) |
| Extent CAL ≥4 mm | 0.255 | 0.047 (-0.016, 0.110) |
| Periodontal sites observed | -0.240 | — |

**Note.** Rank correlations describe the monotonic association between each periodontal measure and HbA1c. Joint age-adjusted coefficients are from a survey-weighted linear regression model including EPE, mean CAL, extent CAL ≥4 mm, linear age, and quadratic age simultaneously. Coefficients represent the difference in HbA1c percentage points per one weighted standard-deviation increase in each periodontal measure, conditional on the other periodontal summaries and age terms. Periodontal sites observed was included in the rank-correlation analysis as an indicator of observability but was not included as a periodontal burden measure in the joint model. Negative or attenuated coefficients in the joint model should be interpreted cautiously because EPE, mean CAL, and extent CAL ≥4 mm are correlated and the model estimates conditional rather than total associations. Confidence intervals are design-based and conditional on the fitted periodontal-attribution model and the assigned CAL specification shown in each row. They do not incorporate uncertainty in the external attribution calibration. CAL = clinical attachment loss; EPE = Expected Periodontitis Experience.

### Supplementary Appendix C. Sensitivity to the minimum assigned CAL value

The primary EPE specification assigned missing tooth positions an advanced CAL value equal to the greater of 10 mm or the participant’s maximum observed tooth-level CAL. This rule was intended to provide a conservative advanced-destruction value for the expected missing-tooth component, not to recover the true pre-extraction CAL value of any individual missing tooth. Because the selected 10-mm value is an analytic assumption, we examined whether the main findings were sensitive to this parameter.

Sensitivity analyses recalculated EPE after replacing the 10-mm floor with alternative minimum assigned CAL values of 6, 8, 12, and 14 mm, while retaining the participant-specific maximum rule. Thus, for each specification, the assigned advanced CAL value for missing teeth was defined as the greater of the specified floor value or the participant’s maximum observed tooth-level CAL. For each recalculated EPE measure, we repeated the EPE component summaries, HbA1c association models, design-weighted rank correlations, and late-life curvature analyses. These analyses were used to evaluate robustness to the missing-tooth burden parameter rather than to select an optimal value.

As expected, increasing the minimum assigned CAL value increased mean EPE and the expected missing-tooth component. Across the sensitivity range, EPE increased from 2.13 under the 6-mm specification to 2.45 under the 14-mm specification, and the expected missing-tooth component increased from 0.30 to 0.63 on the mean-tooth scale (Supplementary Table S6). The primary 10-mm specification remained close to the center of this range.

The external-alignment results were also stable across specifications (Supplementary Table S7). The age-adjusted HbA1c coefficient for EPE increased gradually from 0.123 under the 6-mm specification to 0.154 under the 14-mm specification, and the design-weighted rank correlation with HbA1c increased from 0.256 to 0.307. This monotonic increase is expected because higher assigned missing-tooth burden gives greater weight to accumulated tooth loss.

Late-life curvature showed the same qualitative pattern across the sensitivity range (Supplementary Table S8; Supplementary Figure S6). Among adults with normal glycaemia and prediabetes, EPE showed positive late-life curvature across all assigned CAL values, although confidence intervals included zero for most prediabetes estimates. Among adults with diabetes, EPE showed modest negative curvature across all specifications, ranging from −0.069 (under the 6mm model) to −0.047 (under the 12mm model), with all confidence intervals including zero. These estimates remained less negative than the corresponding curvature for conventional observed-site summaries reported in Supplementary Table S4.

Overall, the sensitivity analysis indicates that the main conclusions were not driven by the specific 10-mm assigned CAL floor. Higher assigned CAL floors modestly strengthened the EPE-HbA1c association (Figure S5) and increased EPE curvature among adults without diabetes (Figure S6), but the substantive interpretation was unchanged: EPE retained cumulative age-patterning and external alignment across plausible values for the assigned missing-tooth CAL parameter.

**Supplementary Table S6.**

*Sensitivity of EPE components to the minimum assigned CAL value for missing tooth positions*

| **CAL floor (mm)** | **Mean assigned missing CAL (mm)** | **Expected missing- tooth component** | **EPE, mean-tooth scale** |
| --- | --- | --- | --- |
| 6 | 6.27 (6.24, 6.29) | 0.30 (0.28, 0.32) | 2.13 (2.08, 2.17) |
| 8 | 8.09 (8.08, 8.10) | 0.37 (0.35, 0.39) | 2.20 (2.15, 2.24) |
| 10 | 10.02 (10.02, 10.03) | 0.45 (0.43, 0.48) | 2.28 (2.23, 2.33) |
| 12 | 12.01 (12.00, 12.01) | 0.54 (0.51, 0.57) | 2.37 (2.31, 2.42) |
| 14 | 14.00 (14.00, 14.00) | 0.63 (0.59, 0.66) | 2.45 (2.40, 2.51) |

**Note**. Confidence intervals are design-based and conditional on the fitted periodontal-attribution model and the assigned CAL specification shown in each row. They do not incorporate uncertainty in the external attribution calibration. The primary specification used a 10-mm minimum assigned CAL value. The assigned missing CAL value was defined as the greater of the specified floor value or the participant’s maximum observed tooth-level CAL. The expected missing-tooth component is expressed on the same mean-tooth scale as EPE.

CAL = clinical attachment loss; EPE = Expected Periodontitis Experience.

**Supplementary Table S7.**

*Sensitivity of HbA1c association estimates to the minimum assigned CAL value for missing tooth positions.*

| **CAL floor (mm)** | **HbA1c beta, unadjusted** | **HbA1c beta, age-adjusted** | **Rank correlation with HbA1c** |
| --- | --- | --- | --- |
| 6 | 0.176 (0.145, 0.206) | 0.123 (0.093, 0.153) | 0.256 |
| 8 | 0.189 (0.159, 0.219) | 0.133 (0.102, 0.164) | 0.277 |
| 10 | 0.201 (0.171, 0.230) | 0.143 (0.112, 0.174) | 0.291 |
| 12 | 0.208 (0.179, 0.237) | 0.150 (0.118, 0.182) | 0.300 |
| 14 | 0.212 (0.184, 0.241) | 0.154 (0.122, 0.186) | 0.307 |

**Note**. HbA1c beta values are survey-weighted linear regression coefficients with 95% confidence intervals and represent the difference in HbA1c percentage points per one weighted standard-deviation increase in EPE. Age-adjusted models included linear and quadratic age terms. Rank correlations are design-weighted rank correlations between EPE and HbA1c. CAL = clinical attachment loss; EPE = Expected Periodontitis Experience.

**Supplementary Table S8.**

*Sensitivity of EPE late-life change and curvature to the minimum assigned CAL value for missing tooth positions*

| **CAL floor (mm)** | **Glycemic status** | **Fitted change, 60–70 years** | **Fitted change, 70–80 years** | **Late-life curvature** |
| --- | --- | --- | --- | --- |
| 6 | Normal | 0.241 (0.203, 0.279) | 0.266 (0.204, 0.328) | 0.025 (-0.002, 0.051) |
| 6 | Prediabetes | 0.182 (0.132, 0.231) | 0.202 (0.114, 0.289) | 0.020 (-0.022, 0.062) |
| 6 | Diabetes | 0.094 (0.029, 0.159) | 0.025 (-0.107, 0.157) | -0.069 (-0.143, 0.005) |
| 8 | Normal | 0.297 (0.259, 0.336) | 0.340 (0.278, 0.401) | 0.042 (0.017, 0.068) |
| 8 | Prediabetes | 0.232 (0.182, 0.282) | 0.260 (0.172, 0.349) | 0.028 (-0.015, 0.071) |
| 8 | Diabetes | 0.156 (0.093, 0.219) | 0.100 (-0.030, 0.229) | -0.056 (-0.129, 0.017) |
| 10 | Normal | 0.360 (0.318, 0.401) | 0.420 (0.355, 0.485) | 0.060 (0.034, 0.086) |
| 10 | Prediabetes | 0.293 (0.239, 0.346) | 0.331 (0.236, 0.425) | 0.038 (-0.008, 0.084) |
| 10 | Diabetes | 0.223 (0.156, 0.291) | 0.174 (0.036, 0.312) | -0.049 (-0.126, 0.028) |
| 12 | Normal | 0.426 (0.378, 0.473) | 0.504 (0.432, 0.576) | 0.078 (0.051, 0.106) |
| 12 | Prediabetes | 0.355 (0.297, 0.414) | 0.403 (0.299, 0.507) | 0.047 (-0.003, 0.097) |
| 12 | Diabetes | 0.291 (0.216, 0.367) | 0.244 (0.092, 0.396) | -0.047 (-0.131, 0.036) |
| 14 | Normal | 0.494 (0.440, 0.548) | 0.591 (0.511, 0.671) | 0.097 (0.067, 0.127) |
| 14 | Prediabetes | 0.419 (0.354, 0.484) | 0.475 (0.360, 0.590) | 0.056 (0.001, 0.111) |
| 14 | Diabetes | 0.360 (0.275, 0.444) | 0.313 (0.145, 0.481) | -0.047 (-0.138, 0.044) |

**Note**. Values are fitted changes with 95% confidence intervals in parentheses from survey-weighted quadratic age models. Late-life curvature was calculated as the fitted change from ages 70–80 years minus the fitted change from ages 60–70 years. Positive values indicate a larger fitted increase from ages 70–80 years; negative values indicate attenuation in later life. Confidence intervals are design-based and conditional on the fitted periodontal-attribution model and the assigned CAL specification shown. They do not incorporate uncertainty in the external attribution calibration. CAL = clinical attachment loss; EPE = Expected Periodontitis Experience.

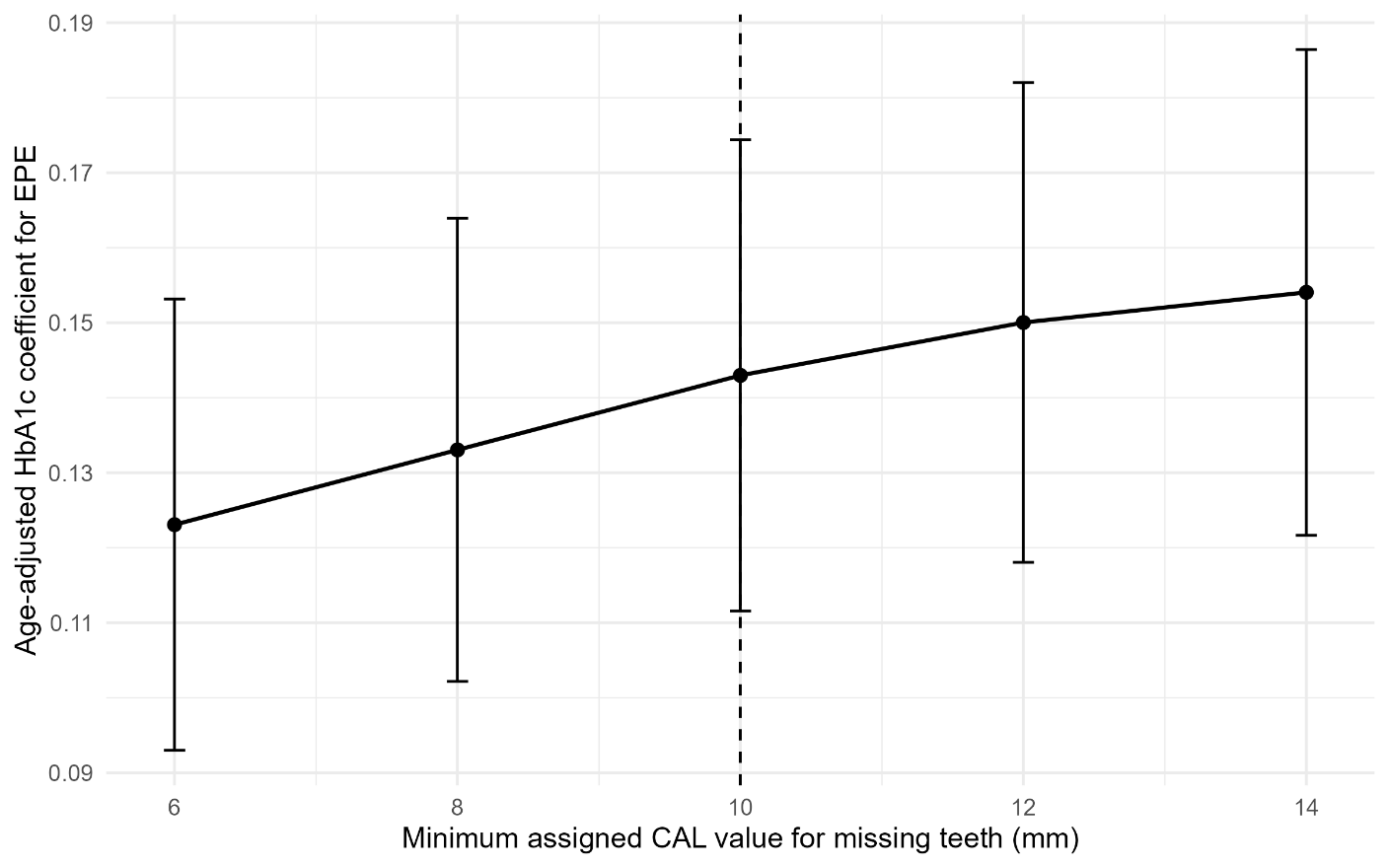

**Supplementary Figure S5. Sensitivity of the age-adjusted EPE-HbA1c association to the minimum assigned CAL value for missing tooth positions.** Points show survey-weighted age-adjusted HbA1c coefficients for EPE. 95% design-based confidence intervals conditional on the fitted periodontal-attribution model and the indicated assigned CAL specification. The vertical dashed line indicates the primary 10-mm specification.

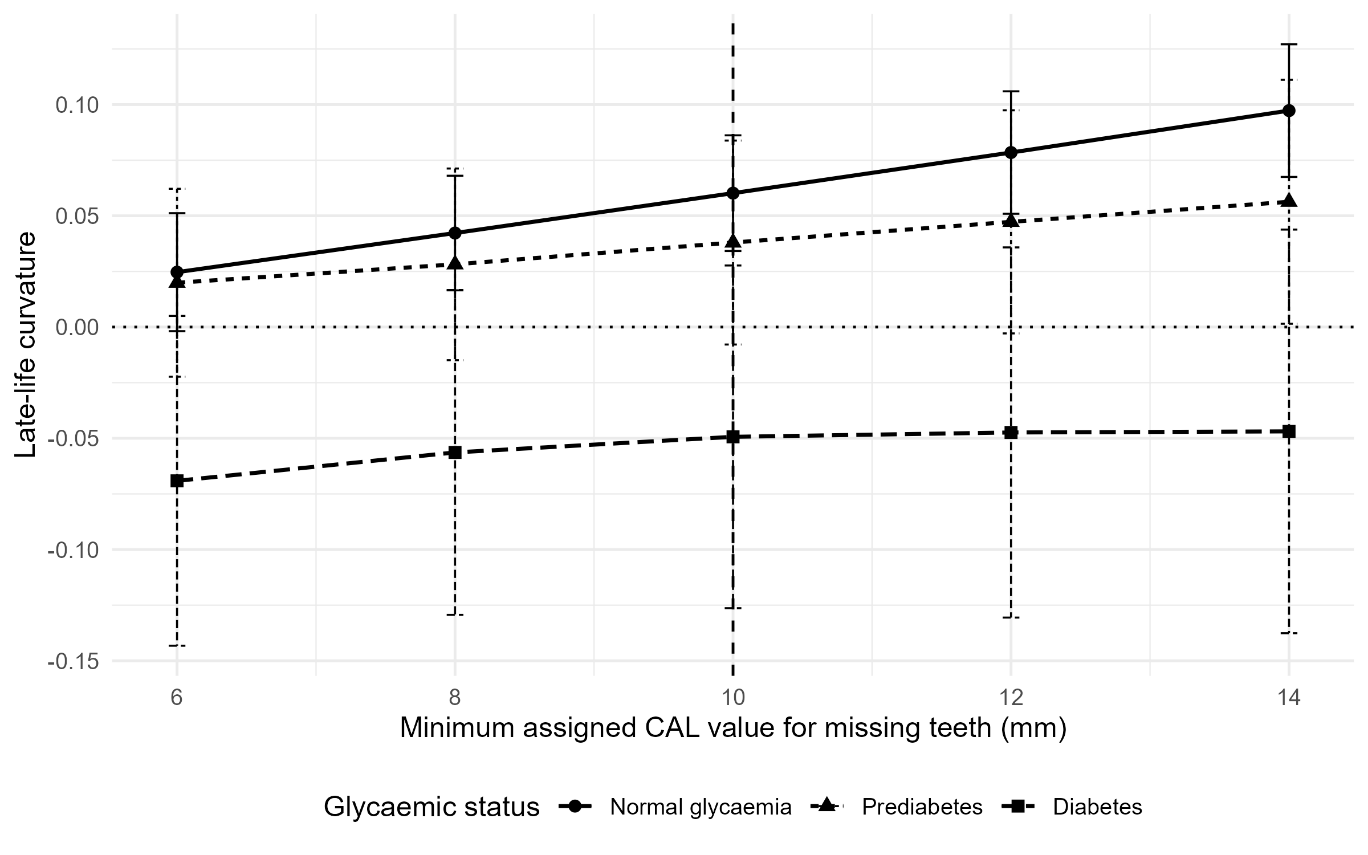

**Supplementary Figure S6. Sensitivity of EPE late-life curvature to the minimum assigned CAL value for missing tooth positions, by glycaemia status.** Late-life curvature was calculated as the fitted change from ages 70-80 years minus the fitted change from ages 60-70 years. The horizontal dotted line indicates zero curvature, and the vertical dashed line indicates the primary 10-mm specification. 95% design-based confidence intervals conditional on the fitted periodontal-attribution model and the indicated assigned CAL specification.
